# Integrating estimation of eye health indicators into trachoma prevalence surveys: a feasibility study in Ethiopia and Nigeria

**DOI:** 10.64898/2026.07.20.26358377

**Authors:** Xiaotong Han, Chrissy h Roberts, Oluwatosin B Adekeye, Jafer Kedir, Zerihun Kura, David Adekunle, Vera Lucia Alves Carneiro, Tesfahun Bishaw, Sarah Boyd, Michael Dejene, Alemu Gemechu, Cristina Jimenez, Fikreab Kebede, Marzieh Katibeh, Caleb Mpyet, Nicholas Olobio, Chris Ostendorf, Anthony W. Solomon, Emawayish Taye, Beatrice Varga, Melissa Witte, Stuart Keel, Emma M. Harding-Esch

## Abstract

**Background:** To evaluate the feasibility of generating data for eye health indicators (EHI), including effective refractive error coverage (eREC) and prevalence of vision impairment, in conjunction with trachoma prevalence surveys.

**Methods:** All individuals aged ≥6 years participating in trachoma prevalence surveys in two evaluation units (EUs) in each of Nigeria and Ethiopia were invited to participate. In parallel with the routine trachoma survey, a separate team assessed EHI metrics under two visual acuity (VA) measurement protocols. In Nigeria, one EU received the standard VA protocol, while the other received a simplified VA protocol. In Ethiopia, participants were randomised for protocol allocation. Acceptability and feasibility were assessed based on fieldwork cost and time and in-depth interviews with key stakeholders.

**Findings:** There were 3806 participants in Nigeria and 4259 in Ethiopia. Prevalence of distance and near vision impairment was 8·4% and 55·3% in Nigeria, and 6·2% and 54·2% in Ethiopia. Distance eREC was 0·0% (95%CI 0·0-0·0) in Nigeria and 1·2% (0·0-3·5) in Ethiopia under the standard protocol, and 0·9% (0·0-2·7) and 9·1% (0·6-17·7) under the simplified protocol. Near eREC was 0·4% (0·0-1·2) (standard) and 1·2% (0·0-2·5) (simplified) in Nigeria, and 2·1% (0·0-4·6) (standard) and 0·9% (0·0-2·1) (simplified) in Ethiopia. The integrated approach was more time- and cost-efficient compared to conducting separate trachoma and EHI surveys. Interviews indicated broad support among stakeholders for the integrated approach.

**Interpretation:** There was a large unmet refractive correction need in the participating communities. Integrating EHI measurement into trachoma surveys was acceptable and feasible.

**Funding:** Christian Blind Mission, The Carter Center, Sightsavers and Fred Hollows Foundation.

## Introduction

Vision impairment (VI) remains a major global public health problem. Globally, it is estimated that over 2·2 billion people have VI and, of which approximately 1 billion could have been prevented or has yet to be addressed.^1^ The burden of VI is projected to increase due to an aging population, alongside projected increases in myopia in the younger population due to lifestyle-related risk factors.^2^

Robust data are the backbone to solving any public health problem, supporting targeted intervention, assessment of intervention effectiveness and efficient resource allocation to areas of greatest need.^3^ The World Health Organization (WHO) has developed the Eye Care Indicator Menu to support monitoring and actions of eye care at national and subnational level.^4^ Included within this menu are population-based eye health indicators (EHI), including prevalence of distance and near VI, effective refractive error coverage (eREC), and effective cataract surgical coverage (eCSC). eREC, defined as the proportion of people in need of refractive error correction who have received services (spectacles, contact lenses, or refractive surgery) and have a good quality outcome, has been listed into WHO’s results framework of the 14th General Programme of Work.^5–7^ In May 2021, at the Seventy-fourth World Health Assembly, WHO Member States endorsed a global target of achieving a 40% increase in eREC by 2030, while recommending that countries with a baseline eREC of ≥60% should strive for universal coverage.^8^

Globally, there remain several challenges with producing robust estimates of priority population-based EHIs, including eREC. Firstly, there is a paucity of data; for example, global estimates of eREC are derived from only approximately 70 countries, with data sometimes dating back 20 years.^6^ Second, there are few data on populations aged <50 years (an age-group in which refractive error is common) and in certain geographical areas, particularly in high-income countries and the European and Americas regions.^6,9–11^ Third, the data are frequently sub-national in representation and not systematically collected, with location largely driven by the presence of non-governmental organisations’ eye care programmes. When these challenges are considered together with the current fiscal environment, it is clear that new and cost-effective solutions are needed to strengthen epidemiological data for eye health.

Tropical Data supports health ministries worldwide to conduct standardised, epidemiologically robust trachoma prevalence surveys that comply with WHO recommendations and with quality assurance and control at each step of the survey process.^12^ By January 2026, Tropical Data had supported surveys in over 4000 Evaluation Units (EUs, generally equivalent to districts), across 55 countries, examining over 13 million people. Integrating population-based EHI data into Tropical Data surveys offers a unique opportunity to leverage surveys already focusing on eyes, together with Tropical Data’s existing methodological, technological and logistical infrastructure, and its wide geographic reach. This study aimed to evaluate the acceptability and feasibility of integrating EHI data collection within routine trachoma surveys supported by Tropical Data, assessing (1) EHI estimates in two EUs in each of Ethiopia and Nigeria; (2) the cost and time of the integrated approach compared to standalone trachoma surveys; and (3) stakeholder feedback through in-depth interviews (IDIs).

## Methods

### Ethics Statement

This study was conducted in accordance with the Helsinki Declaration. Ethical approval was obtained from the London School of Hygiene & Tropical Medicine (LSHTM) Ethics Committee (Ref 16692), the National Health Research Ethics Committee of Nigeria (NHREC/01/01/2007) and the Regional State Health Bureau ethical clearance committee in Ethiopia (BF0HHQ1038/H). Verbal consent for participation was obtained from individuals, or from the parent or legal guardian in the case of those aged <18 years. Consent was documented in an Android app before collecting any data. For the qualitative study, participants provided written informed consent, which also included consent for audio recording of the interview. The study is reported following the Strengthening the Reporting of Observational Studies in Epidemiology (STROBE) reporting guidelines (appendix 1).^13^

### Trachoma surveys

According to the WHO-recommended methodology for trachoma population-based prevalence surveys,^14^ estimates should be generated at EU level, the “normal administrative unit for health care management, consisting of a population unit between 100,000 and 250,000 persons”. Per EU, 20-30 clusters are sampled with probability of selection proportional to population size, and all consenting residents aged ≥1 year in randomly selected households per cluster.^15^ The number of households per cluster is determined, in advance for the EU, by how many a team can reasonably survey in one day of fieldwork.

### Study sites

In Nigeria, EUs correspond with local government areas, and in Ethiopia, EUs correspond with woredas. The study took place in 2 EUs (60 clusters) in each country, between 30^th^ June and 14^th^ July 2024 in Nigeria, and between 28^th^ October and 2^nd^ November 2024 in Ethiopia (Figure 1).

**Figure 1.**
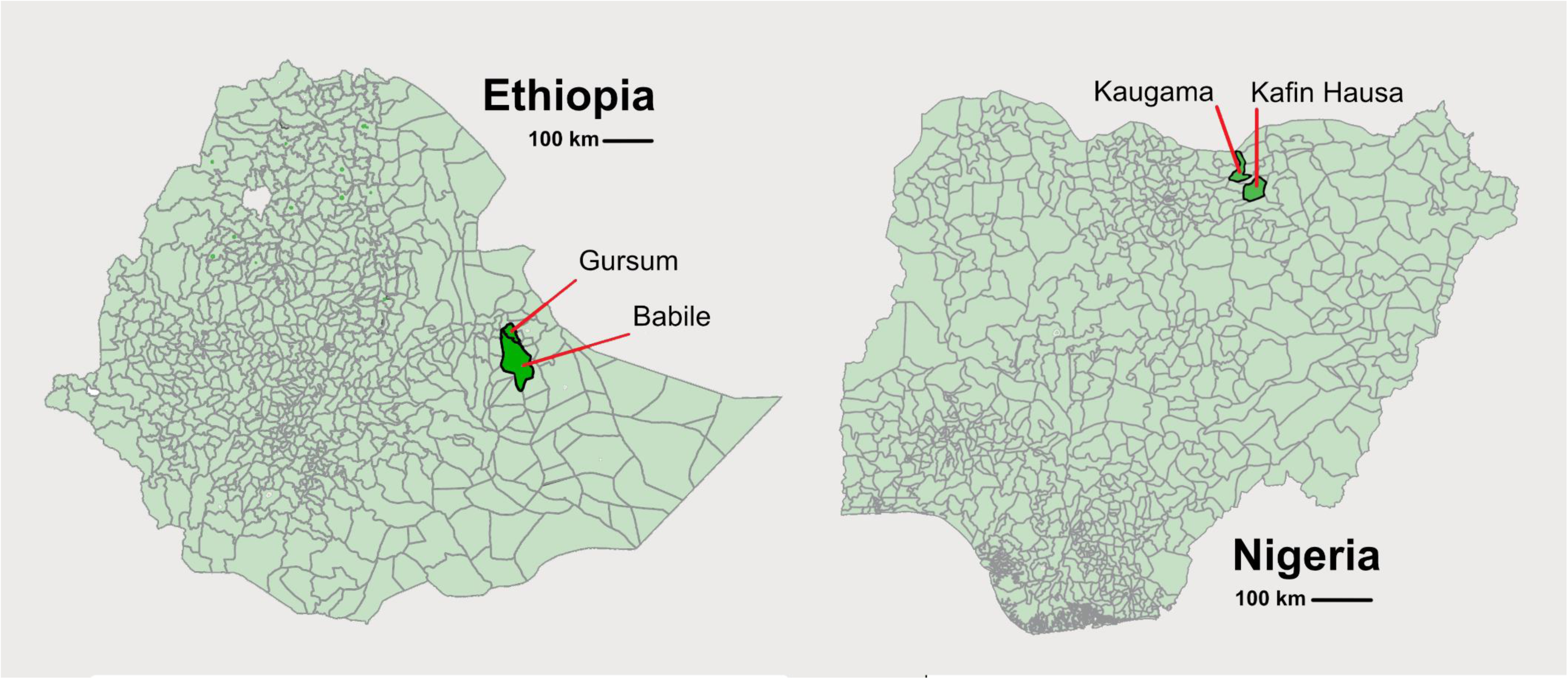
Map of study recruitment evaluation units (EUs) in Ethiopia and Nigeria. Dark green indicates the EUs enrolled from Ethiopia and Nigeria in the study. The boundaries and names shown and the designations used on this map do not imply the expression of any opinion whatsoever on the part of the authors, or of the institutions with which they are affiliated, concerning the legal status of any country, territory, city or area or of its authorities, or concerning the delimitation of its frontiers or boundaries. Dotted lines on maps represent approximate border lines for which there may not yet be full agreement.

### Sample size calculation

The sample size calculation was based on that recommended by WHO for trachoma prevalence surveys, resulting in a target of 1164 children aged 1-9 years to enumerate per EU.^14^

### Training

Trachoma survey graders and recorders were certified according to the standard Tropical Data training system.^15^ EHI examiners received a 2-day training on visual acuity (VA) measurement using the Peek Acuity application.^16^ Field coordination between the trachoma and EHI teams was also rehearsed during training.

### Data collection

For trachoma, the standard Tropical Data methodology was employed (appendix 2, supplementary figure 1A).^14,17^ For EHI, the examiner worked in parallel to the trachoma team (appendix 2, supplementary figure 1B). The VA assessment workflow is shown in Figure 2. All individuals aged ≥6 years selected for inclusion in the trachoma surveys were invited to participate in EHI data collection. Data were collected on Android smartphones using ODK.^18^ For VA assessment, a Peek Acuity prototype was used. The VA testing flow was pre-set in the data collection app, allowing automatic switching between the ODK and Peek apps. As part of the feasibility assessment, two VA protocols were employed: the standard protocol, assessing VA at four different cut-offs (6/12, 6/18, 6/60, and 3/60), and the simplified protocol that only determined whether VA was ≥6/12. In Nigeria, one EU was assessed using the standard protocol and the other EU using the simplified protocol. In Ethiopia, to avoid potential confounding of results because of intrinsic differences between EUs, participants were randomised to receiving the standard or simplified protocol. In Nigeria, both near and distance VA were measured, one eye at a time (monocular). Given that presbyopia usually affects both eyes similarly, in Ethiopia, the faster approach of measuring near VA, was adopted.^19^

**Figure 2.**
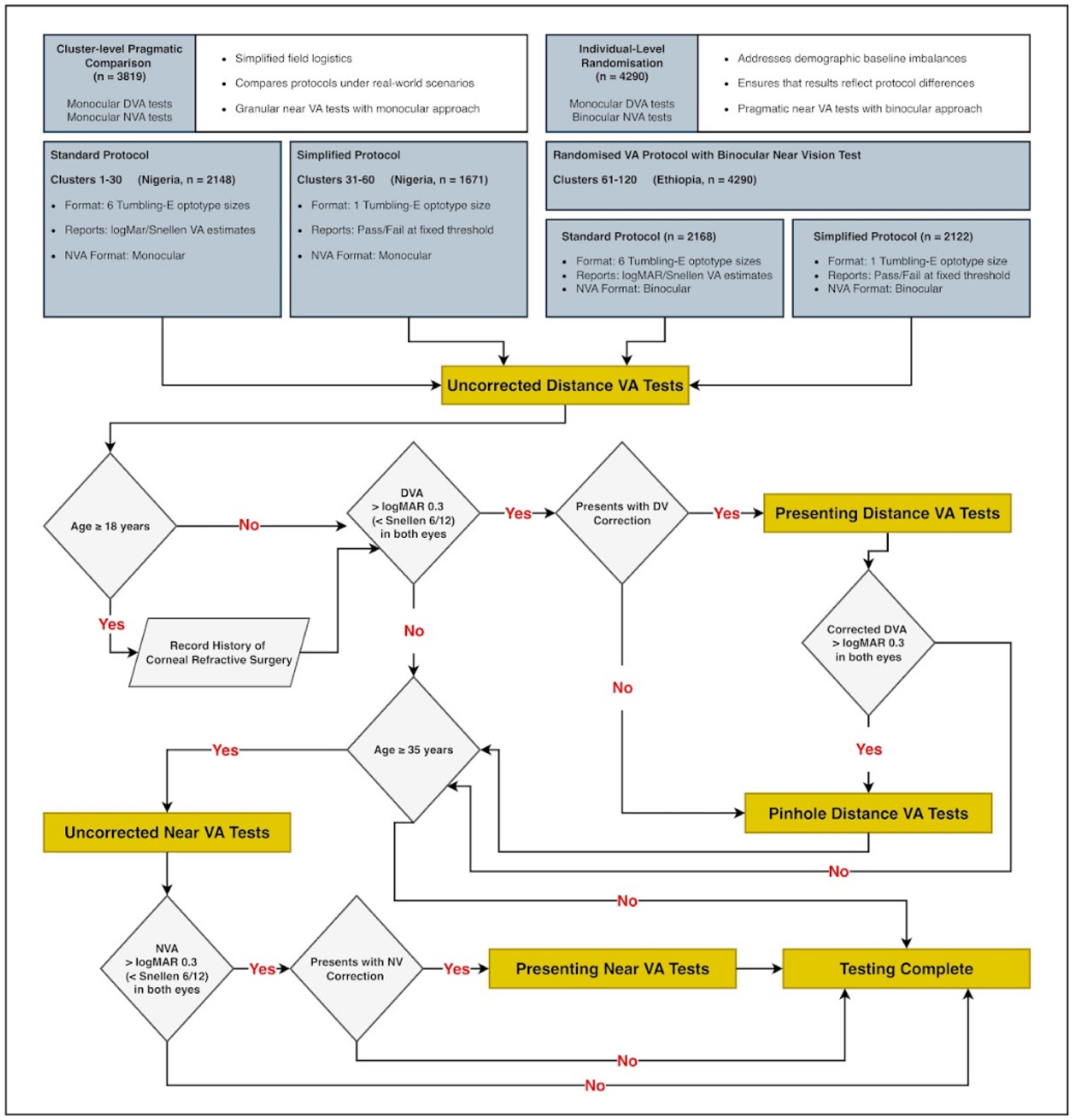
Visual acuity assessment workflow. A visual abstract of the study protocol, showing the sequence of visual acuity assessments applied to participants, including uncorrected, presenting, and pinhole distance vision tests, with additional near vision testing for those aged ≥35 years. It also illustrates how standard and simplified protocols are implemented across different study components. Abbreviations: DVA=distance visual acuity, NVA=near vision acuity; DV=distance vision; NV=near vision.

Individuals identified with active trachoma were offered 1% tetracycline eye ointment to apply at home twice daily for 6 weeks, while those with trichiasis were referred to the nearest health facility designated to provide trachomatous trichiasis surgery. All individuals identified with VI were informed of the results and referred to the nearest health facility for detailed ophthalmic examination.

### Cost and time data collection

Cost data for the 4 EUs’ surveys were provided by the national programmes and their implementing partners in Excel spreadsheets, using an ingredients approach, including cost of training, transportation, consumables, salaries and allowances. A narrative of survey implementation in the field was also provided. To calculate the extra time VA measurement added, data were collected using the automatically collected time stamps in the Android smartphones on total time in the field per cluster for the trachoma team, and number of minutes per person to do the VA measurement.

### Stakeholder feedback data collection

Semi-structured IDIs were held with key international, national, and local level stakeholders. Interviews were conducted either in English or the local language, as appropriate, in a private room or via phone or video link (e.g., Zoom) by experienced qualitative researchers. IDIs followed a topic guide (appendix 3) to assess participants’ experience and views on opportunities and challenges of integrating EHI measurement into trachoma surveys. Individuals involved in the fieldwork were also asked what went well during the project, and what could be improved.

### Data management and analysis

VA data were automatically encrypted by the data collection app, uploaded, and stored on LSHTM institutional servers. VA data analysis was conducted independently by two statisticians, one using R (version 4·5·1)^20^ and the other using STATA 16·0 (StataCorp, College Station TX, USA). EHI were calculated in broad alignment with Rapid Assessment of Avoidable Blindness (RAAB) protocols^3,21^, using WHO definitions (appendix 4). Indicators were derived from individual-level VA measurements and expressed as population-level proportions using predefined numerator and denominator sets, with cluster-adjusted estimates. Corresponding 95% confidence intervals (CIs) were derived using design-based methods. Communities were defined as the primary sampling unit, and age-sex post-stratification was applied to align the sample with the underlying population distribution. Recent national censuses were unavailable for both Nigeria and Ethiopia, so age-sex population distributions were derived from modelled estimates based on the most recent national census data. Models for the populations of Nigeria (circa 2022) and Ethiopia (circa 2023) were obtained from the Humanitarian Data Exchange (HDx). Protocol deviations were quantified as the proportion of participants for whom the observed testing sequence did not conform to the predefined protocol logic. EHI were additionally estimated with stratification by sex and age group using the same predefined numerator and denominator sets, with design-based methods applied to account for the cluster sampling structure.

Distance VI was defined as bilateral presenting VA <6/12, with pinhole VA used to assess potential improvement. Near VI was defined as bilateral presenting VA <6/12 among participants aged ≥35 years. Uncorrected distance VI was defined as bilateral uncorrected VA <6/12. Pinhole VI was defined as bilateral VA <6/12 following pinhole correction. Correctable distance VI was defined as bilateral presenting VI that improved to VA ≥6/12 with pinhole and was expressed as the proportion of individuals with VI meeting this criterion. Near correctable VI was estimated using a proxy approach, defined as the proportion of individuals with presenting near VI (aged ≥35 years) assumed to be correctable in the absence of measured pinhole correction. The relative quality gap was calculated as the percentage difference between REC and eREC relative to REC.

Survey time and costs were calculated for the following three scenarios: standalone trachoma surveys; integrated survey “separate” approach, where trachoma and VA data recorders would work in parallel (as was done in the current study); and integrated survey “combined” approach, where the trachoma data recorder would also be responsible for measuring VA (appendix 2, supplementary figure 1A-C). For costs, the relative cost of integrating EHI on top of trachoma-only survey costs was calculated by cost category (sensitisation, training, fieldwork). Cost data from the feasibility study were extrapolated to scenarios where teams spent two days per cluster instead of one, or having the integrated survey “combined” approach instead of the “separate” approach. To calculate the extra time VA measurement added to the trachoma survey, the trimmed mean number of minutes to conduct the VA assessment per person was derived from audit log timestamps recorded within the data collection application, using a 5% trimmed mean to reduce the influence of implausible outliers while retaining all observations, with durations exceeding 600 seconds excluded as technical artefacts arising from incomplete form closure. For each country and protocol variant, this value was multiplied by the number of examined residents aged ≥6 years per EU recorded in the trachoma dataset.^22^ The median time per cluster within each EU for each scenario was then calculated by adding the extra VA time to the total data collection time for the trachoma team.

IDIs were audio-recorded and transcribed for analysis. Thematic analysis was conducted by experienced researchers from Nigeria and Ethiopia, who independently identified themes, cross-checked them, and reached a consensus.^23^

## Results

### Participant overview

Table 1 presents the demographic characteristics of the samples as unweighted sample counts alongside proportions estimated using survey methods that account for the cluster sampling design, in-line with RAAB protocols. A total of 3819 participants from Nigeria and 4290 from Ethiopia were recruited. Following exclusion due to protocol deviations (appendix 2, supplementary Table 1), the sample sizes for Nigeria were 1664 (standard protocol) and 2142 (simplified protocol), and for Ethiopia were 2149 (standard protocol) and 2110 (simplified protocol). In Nigeria, participants who received the standard protocol were younger (P=0·011) and had lower educational levels (P<0·001) compared to those who received the simplified protocol. In Ethiopia, due to the use of individual-level randomisation, there was no evidence of demographic differences between participants in the two protocols (Table 1).

**Table 1.**
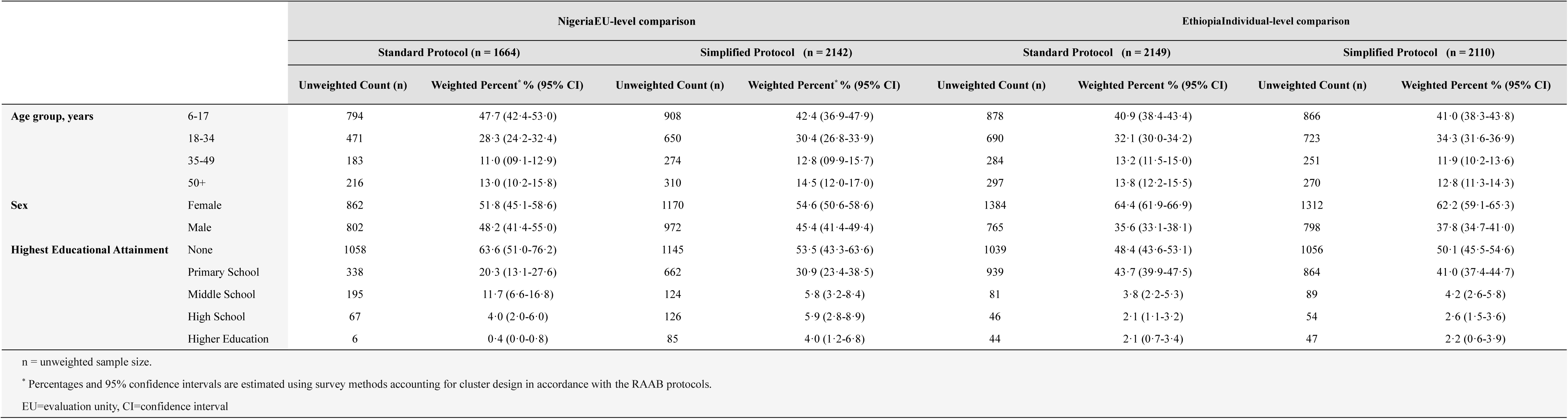
Participant characteristics from cluster-based surveys in Nigeria and Ethiopia.

### EHI

A detailed summary of the various EHI estimates is provided (Table 2). Distance VI prevalence was similar across countries and protocols (Nigeria: 6·7 to 8·4%; Ethiopia: 6·0% to 6·2%). The simplified protocol produced lower estimates of VI based on pinhole VA compared to the standard protocol in Nigeria (1·4% vs 4·2%), but were comparable in Ethiopia (3·7%). Estimates of correctable VI were broadly consistent. eREC were approximately 0% to 1% in Nigeria, and approximately 1% to 9% in Ethiopia. Across both countries, near VI prevalence was high (ranging between 53% to 56%), with substantial correctable burden (41% to 52%), but minimal coverage (REC ≤2·4%).

**Table 2.** Population prevalence estimates of eye health indicators in Nigeria and Ethiopia.

| Indicator | Concept | Nigeria Full Protocol | Nigeria Simplified Protocol | Ethiopia Full Protocol | Ethiopia Simplified Protocol |
| --- | --- | --- | --- | --- | --- |
| **Bilateral distance VI (uncorrected VA) (%)** | Underlying Need | 8·4 (5·9-10·9) | 6·7 (5·0-8·3) | 6·2 (5·2-7·2) | 6·0 (5·1-6·9) |
| **Bilateral distance VI (presenting VA) (%)** | Current Provision | 8·4 (5·9-10·9) | 6·6 (4·9-8·3) | 6·2 (5·2-7·2) | 5·9 (5·0-6·8) |
| **Bilateral distance VI (pinhole VA) (%)** | Potential Improvement | 4·2 (2·6-5·7) | 1·4 (0·9-2·0) | 3·7 (3·0-4·5) | 3·7 (2·9-4·5) |
| **Bilateral distance correctable VI (%)** | Correctable Burden | 4·2 (3·1-5·8) | 5·2 (3·9-6·9) | 2·5 (1·9-3·2) | 2·2 (1·6-3·0) |
| **Distance vision REC (%)** | Access to Correction | 0·0 (0·0-0·0) | 0·89 (0·0-2·68) | 1·2 (0·0-3·5) | 10·6 (1·6-19·5) |
| **Distance vision eREC (%)** | Effective Access to Correction | 0·0 (0·0-0·0) | 0·89 (0·0-2·68) | 1·2 (0·0-3·5) | 9·1 (0·6-17·7) |
| **Distance vision relative quality gap (% of REC) (%)** | Performance Shortfall | NA* | 0·0 (NA<sup>†</sup>) | 0·0 (NA<sup>†</sup>) | 13·7 (NA<sup>†</sup>) |
| **Bilateral near VI (age ≥35) (%)** | Underlying Need | 55·3 (49·6-61·1) | 56·1 (50·6-61·6) | 54·2 (48·3-60·0) | 53·1 (47·2-59·1) |
| **Bilateral near correctable VI (Proxy; age ≥35)\* (%)** | Correctable Burden | 44·9 (39·0-51·0) | 52·4 (46·5-58·3) | 44·2 (37·9-50·7) | 41·3 (35·7-47·1) |
| **Near vision REC (age ≥35) (%)** | Access to Correction | 0·8 (0·0-1·9) | 2·0 (0·3-3·7) | 2·4 (0·0-4·9) | 1·6 (0·0-3·2) |
| **Near vision eREC (age ≥35) (%)** | Effective Access to Correction | 0·4 (0·0-1·2) | 1·2 (0·0-2·5) | 2·1 (0·0-4·6) | 0·9 (0·0-2·1) |
| **Near vision relative quality gap (% of REC) (%)** | Performance Shortfall | 49·4 (NA<sup>†</sup>) | 41·9 (NA<sup>†</sup>) | 10·5 (NA<sup>†</sup>) | 44·5 (NA<sup>†</sup>) |

\* Quality gap cannot be calculated when REC and eREC are zero.
† Quality gap is a derived difference between two survey estimates, and a valid confidence interval would require propagation of variance within the survey design. Since this is not performed, only the point estimate is reported.
‡ Near vision correctable VI is estimated using a proxy definition because pinhole testing was not performed for near vision in this study. Individuals with uncorrected near visual impairment were classified as correctable if distance visual acuity was ≥6/12 in at least one eye.
Abbreviations: CI=confidence interval, REC= refractive error coverage, eREC=effective refractive error coverage, VI=vision impairment, VA=visual acuity.

Age- and sex-stratified estimates (appendix 2, supplementary figures 2-5) showed patterns consistent with the overall results. Distance VI increased with age and was highest in older age groups across both protocols and countries. Near VI was higher in the oldest age group. Sex-stratified estimates were broadly similar across males and females, with no consistent differences across indicators. Patterns were comparable between standard and simplified protocols within each country.

### Cost and time of VA assessment

If fieldwork could be completed in one day per cluster (the usual trachoma methodology), integrating EHI added one-third to the trachoma-only survey costs in Nigeria and 60% in Ethiopia, regardless of approach used (“combined” or “separate”) (Table 3). Integration led to cost-savings compared to doing separate trachoma and EHI surveys, for example, through combined advocacy costs, reduced trainer logistics (e.g., shared travel costs to training venues and survey sites), and reduced fieldwork costs (e.g., shared costs of supervisors and local guides).

**Table 3.** Additional relative cost of integrating eye health indicators (EHI) into trachoma prevalence surveys.

| Cost category | Nigeria |  |  | Ethiopia |  |  |
| --- | --- | --- | --- | --- | --- | --- |
|  | Integrated survey – “combined”* |  | Integrated survey – “separate”† | Integrated survey – “combined”* |  | Integrated survey – “separate” † |
|  | One day/cluster | 2 days/cluster |  | One day/cluster | 2 days/cluster |  |
| Sensitisation | 0% | 0% | 0% | 0% | 0% | 0% |
| Training | 83% | 83% | 85% | 89% | 89% | 58% |
| Fieldwork | 0% | 16% | 3% | 0% | 89% | 72% |
| TOTAL | 33% | 40% | 35% | 56% | 89% | 63% |
| *“Combined”: EHI and trachoma data collected on one Android device. Nurse records visual acuity on Peek and then serves as data recorder for trachoma outcomes. |  |  |  |  |  |  |
| † “Separate”: Nurse for EHI & Peek; data recorder and grader for trachoma, using separate Android devices. This is what was done for this study. Assumes the cluster is surveyed in one day. |  |  |  |  |  |  |

Compared to conducting standalone trachoma surveys, the median data collection time for a “combined” integrated approach survey added about 2·5 hours per cluster in Ethiopia, 2·5 hours in Nigeria using the simplified protocol, and 4·5 hours in Nigeria using the standard protocol (appendix 5, supplementary table 2). The average (5% trimmed mean) time per person to conduct the VA assessment was 1m 18s (Nigeria) or 1m 17s (Ethiopia) for the simplified protocol, and 1m 54s (Nigeria) or 1m 55s (Ethiopia) per person for the standard protocol. A reduction in time required for VA testing with ongoing field work was also observed (appendix 2, supplementary figure 6).

### Acceptability of EHI assessment

We surveyed 26 individuals, including international stakeholders (n=10), national programme coordinators and policymakers (n=6), and field team members (5 in each country). The integrated approach received generally favourable feedback from stakeholders, as it could help community members better understand their vision status, and it enabled resource-sharing, leading to cost-savings and more efficient training (Figure 3). However, challenges were noted, including the lack of immediate interventions, such as eyeglass provision, the need for additional time to coordinate survey planning nd data collection, and securing the funds for the additional components. Most participants preferred the simplified protocol for its ease-of-use, and stakeholders recommended improving training and supervision integration, programmes moving towards a more holistic community eye health approach rather than a disease-specific focus, providing reading glasses when needed, and establishing clearer guidelines for roles in future integrated surveys.

**Figure 3.**
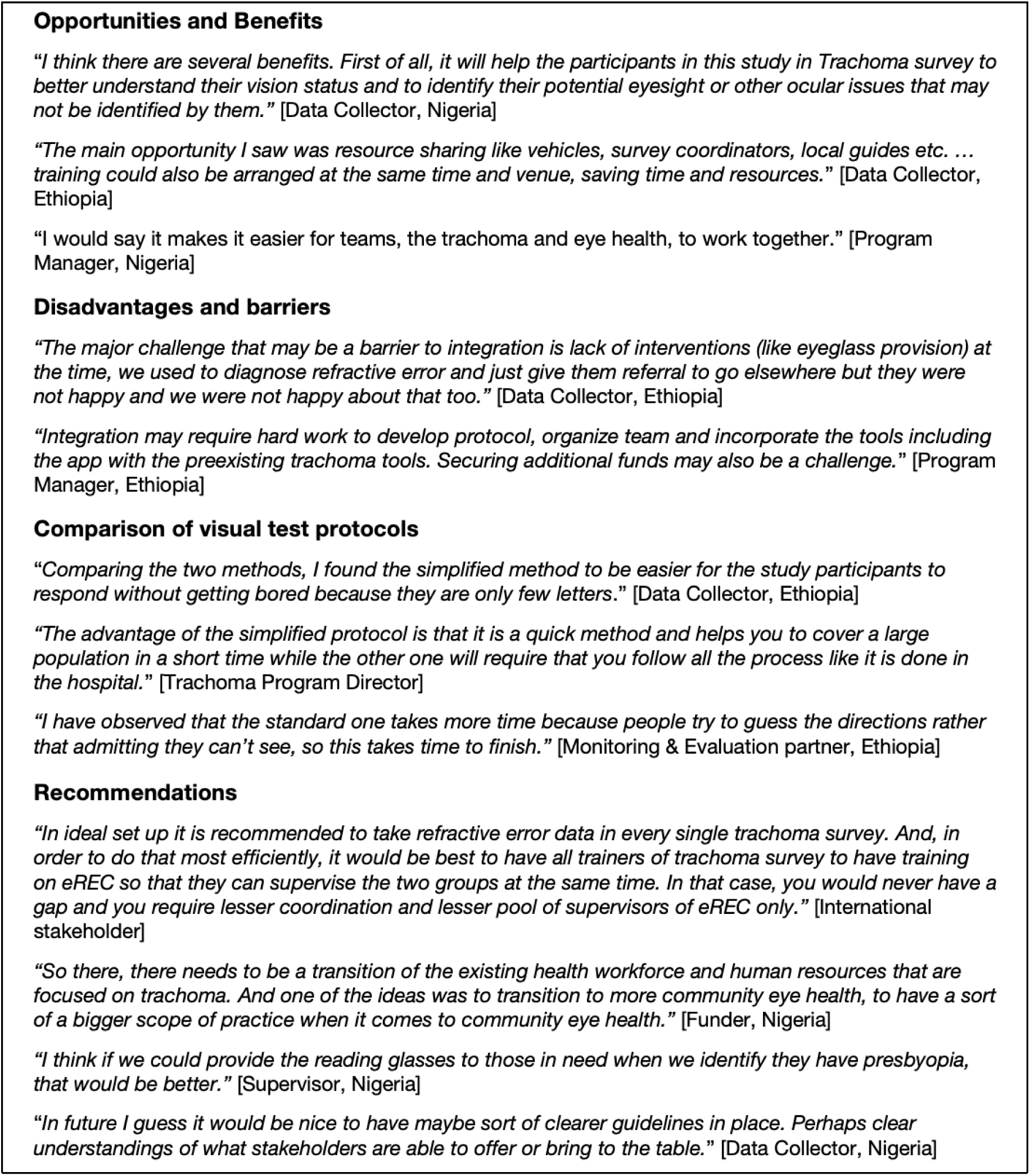
Stakeholder feedback regarding the acceptability and feasibility of integrating eye health indicator assessment in trachoma surveys.

## Discussion

This study provides the first evaluation of integrating population-based EHI assessment within routine trachoma surveys. Our findings demonstrate that an integrated approach is both feasible and acceptable, offering time and cost savings compared to standalone assessments. We also highlight a large unmet need, with less than 2.5% of the survey population in need of eyeglasses having access to them.

Our study revealed low eREC (both distance VI and near VI) amongst the surveyed populations in both Nigeria and Ethiopia. These values are notably lower than recent global estimates for the Sub-Saharan African region (28·3%).^6^ In addition, the prevalence of distance and near VI are among the highest reported globally.^18^ These findings are likely attributed to the rural and remote location of the surveys: we sampled underserved populations that have been infrequently included in previous data collection exercises for EHI. Approximately 40% of distance VI and 75% of near VI cases identified were correctable with eyeglasses, indicating an urgent need for concerted actions to improve access to eye care in these regions.

A critical consideration for the feasibility of the integrated survey approach was whether a cluster could be surveyed within one day, as is standard for standalone trachoma surveys.^12^ Our analyses indicated that this is possible with the “combined” integrated approach if the simplified VA protocol is utilised. If the standard VA protocol is employed, then the completion of a cluster in one day would either depend on local contextual factors, such as the time to travel to the cluster and the set-up in the cluster (e.g., distance between households, and household size), or require a reduction in the number of households sampled per cluster. Furthermore, as teams become familiar with the methods, fieldwork will go faster, although improvements are likely to be finite and to plateau over time (appendix 2, supplementary figure 6). Our study also evaluated the implications of utilising a “separate” team to conduct the EHI assessment in parallel to the trachoma assessment. While costs were higher using this approach, the completion of a survey cluster in a single day was achievable, regardless of the VA protocol (simplified or standard). One advantage of the “separate” approach is that the EHI team could likely complete data collection earlier than the trachoma team, which would offer an ideal time for further follow up and provision of management, such as eyeglasses. For example, assuming the provision of near eyeglasses takes about two minutes per person and, as seen in our study, approximately 50% of those aged ≥35 years had near VI and needed eyeglasses, this equates to roughly 13 people per cluster, or 30 minutes of extra time in the field. Given that 826 million people globally live with near VI from uncorrected presbyopia,^24^ and almost none of the participants who required reading glasses in this study already had them, this window for on-field intervention could be highly valuable, with potential to reduce disability associated with near VI. Future work should explore the most effective and operationally feasible approaches for dispensing eyeglasses within survey settings.

A key consideration for determining the appropriateness of using trachoma surveys as the platform for EHI integration is the routine sample size of trachoma surveys. Using data from the trachoma dataset from this study, the mean sample size of individuals aged ≥6 years in the EUs was 3122. In principle, this scale of sampling is compatible with estimation of the selected EHIs, although achieved precision depends on the survey design, clustering and the underlying prevalence of the indicators. For example, under assumed design parameters, a sample size of 3200 would allow a distance eREC population coverage of 4·9% to be estimated with a margin of error of 0·015 at 80% confidence, while a sample size of 927 would allow a distance VI prevalence of 9% to be estimated, with a margin of error of 0·020 at 95% confidence.

The integrated approach evaluated in this study is in-line with the road map for neglected tropical diseases (NTDs) 2021-2030, which calls for more cross-cutting approaches and multisectoral action,^25^ and offers significant potential benefits at global, regional, and national levels. Between 2025 and 2032, a total of 2062 trachoma surveys are planned across 1269 districts in 35 countries.^26^ Notably, 29 of these countries either have no data, or only outdated data, on EHI. Integration of EHI data collection thereby presents great potential to address key data gaps, particularly in the Americas and African regions, and for near VI estimates among children and working-age populations. Other benefits, as widely agreed by stakeholders during our IDIs, include increased population awareness of eye health and vision status, improved population engagement, enhanced efficiency by measuring multiple indicators in a single survey, and fostering collaboration among different health programmes. As indicated in the IDIs, close coordination between the NTD and eye care sectors is a critical element for facilitating future integrated surveys. Careful planning, training and supervision are also needed: it is important that both teams understand each other’s responsibilities and workflows, disaggregate survey tasks, and conduct practice sessions to facilitate seamless collaboration.^27^ Further, integrated surveys require strategic and political commitments and a move away from disease- and activity-specific funding, which can impede integration if responsibilities lie with different organisations.

Some limitations of this study should be considered. First, the study population was limited to two EUs in each of two African countries, and therefore our findings may not be representative of all EUs globally in which this approach could be applied. Second, we did not have data to conduct a cost comparison with a standalone eye health survey. An area for future investigation could be a comparison of costs with those of RAAB surveys.^21^ Third, we did not assess the “combined” integrated approach, or the feasibility of providing eyeglasses at the time of the survey. Provision of post-screening care is of significant public health and pragmatic importance. In order to abide by the principle of “no survey without service”,^28^ appropriate management should be provided to all participants in need. Due to time and funding constraints, we were regrettably unable to provide free eyeglasses during fieldwork and instead were only able to refer them to the nearest health facility for detailed ophthalmic examination. Provision at the time of examination is likely to improve compliance with eyeglass use.^29^ The feasibility and acceptability of providing eyeglasses during the survey is an area for future work.

Electronic data capture (EDC) has well-established advantages, such as its efficiency, reduced likelihood of data entry errors, reduced risk of data loss, and allowing for rapid review and feedback with field teams.^11^ However, in certain settings, for example, where there is insecurity that puts field teams with phones at risk, conventional paper-form VA charts could be used. Visual acuity testing on smartphones can be implemented through various apps including both Peek^16^ and WHO Eyes^30^**Error! Bookmark not defined.**. However, neither Peek nor WHOeyes currently has a freely and publicly available version that can integrate with major data collection platform such as ODK, posing important limitations for large-scale surveillance programmes.

A concern for the sustainability of this integrated approach may be the target year of 2030 for global trachoma elimination, whilst there will always be a need for eyecare. To this end, Tropical Data has been working to develop its methods and resources to support the collection of additional indicators within routine trachoma surveys to support programmatic decision-making. This flexibility, whilst maintaining data quality and standardisation, helps ensure the future sustainability of support provided to health ministries, potentially even reframing the directionality of the integration. The Tropical Data platform, and this study, can also serve as a template for integrating other health indicators into ongoing surveys.

## Conclusion

There is a large unmet need for eye care services in trachoma-endemic populations. This study found that integrating EHI measurements into routine trachoma prevalence surveys is acceptable, feasible and cost-effective, offering great potential to strengthen local and global epidemiological data on eye health, with subsequent improvements for eyecare advocacy, planning and monitoring.

## Contributors

All authors had final responsibility for the decision to submit the manuscript for publication. XH, ChR, SK, and EHE contributed to the conceptualisation, methodology, formal analysis, and writing of the original draft. DA, TB, SB, MD, AG, CJ, FK, CM, NO, CO, ET, and BV contributed to the methodology, data curation and project administration. MK and MW provided support for the Peek prototype and integration. OA, JK, ZK conducted the qualitative data analysis. VLAC and AWS provided study oversight. EHE, SK, CJ, AWS contributed to funding acquisition. XH, ChR, SK and EHE had access to and verified the data reported in the manuscript. All authors contributed to the review and editing of the manuscript. The corresponding authors had full access to all the data in the study and the final responsibility for the decision to submit the manuscript for publication.

## Acknowledgements

ODK was implemented by LSHTM’s Global Health Analytics Group (odk.lshtm.ac.uk). We are grateful to the field teams and all the study participants. We would also like to thank all the participants of the “Stakeholder meeting on the integration of eye health indicators within trachoma surveys”, October 2025, for their feedback on this approach. For the research components, the World Health Organization (WHO) received funding from Christian Blind Mission, and the London School of Hygiene & Tropical Medicine (LSHTM) received funding from The Carter Center. Sightsavers and The Fred Hollows Foundation provided funding for the eye health indicator fieldwork. Core Tropical Data funding for the trachoma survey components was provided by the International Trachoma Initiative and Sightsavers. ChR was supported by the LSHTM Global Health Analytics Group’s ‘Pay What You Can’ funding scheme, a crowdsourced initiative that facilitates innovative and open methods research for the public good. VLAC, AWS and SK are staff members of WHO.

## Data availability

Requests for access to data used for this analysis can be made by contacting the corresponding author. The source code for the statistical analysis is available at https://github.com/chrissyhroberts/eREC_Tools.

## APPENDIX 1

**STROBE Statement—Checklist of items that should be included in reports of *cross-sectional studies***

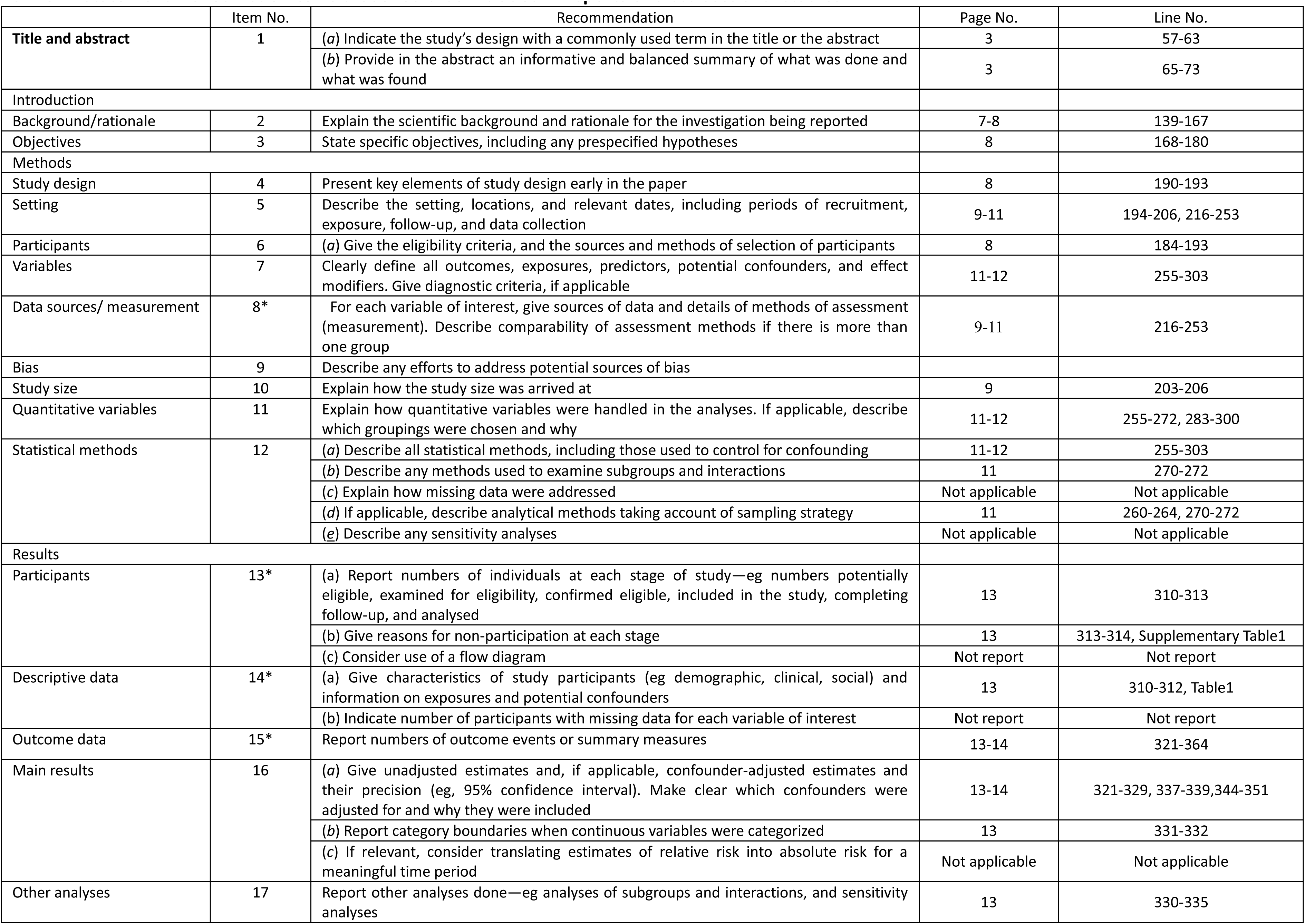
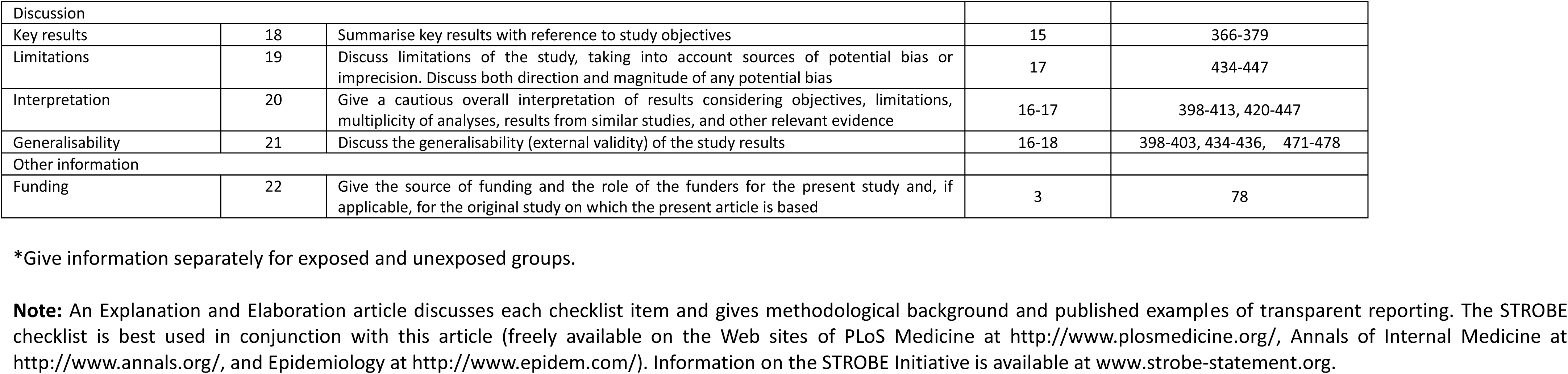

## APPENDIX 2

**Supplementary Table 1.** Number of protocol deviations observed.

| Category | Reason | Nigeria Standard Protocol | Nigeria Simplified Protocol | Ethiopia Standard Protocol | Ethiopia Simplified Protocol |
| --- | --- | --- | --- | --- | --- |
| **No Deviation** | No Deviation | 1664 | 2142 | 2149 | 2110 |
| **Excluded** | Missing uncorrected distance VA | 0 | 1 | 2 | 2 |
|  | Missing presenting distance VA when required | 0 | 0 | 0 | 0 |
|  | Missing pinhole distance VA when required | 6 | 4 | 4 | 1 |
|  | Missing uncorrected near VA when required | 2 | 1 | 13 | 9 |
|  | Missing presenting near VA when required | 0 | 0 | 0 | 0 |
|  | **Any protocol exclusion\* (total unique records removed)** | **7** | **6** | **19** | **12** |
| **Retained but partial†** | Partial uncorrected distance VA (one eye only) | 11 | 22 | 20 | 7 |
|  | Partial presenting distance VA (one eye only) | 0 | 0 | 0 | 0 |
|  | Partial pinhole distance VA (one eye only) | 12 | 5 | 6 | 3 |
| **Audit only-unusual patterns** | Unexpected presenting distance VA | 0 | 0 | 0 | 0 |
|  | Unexpected pinhole distance VA | 0 | 0 | 0 | 0 |
|  | Unexpected presenting near VA | 0 | 0 | 0 | 0 |
| *Any individual may have been excluded because of multiple deviations. †Some partial data relates to individuals with a single testable eye.<br>VA: visual acuity |  |  |  |  |  |

**Supplementary Figure 1.**
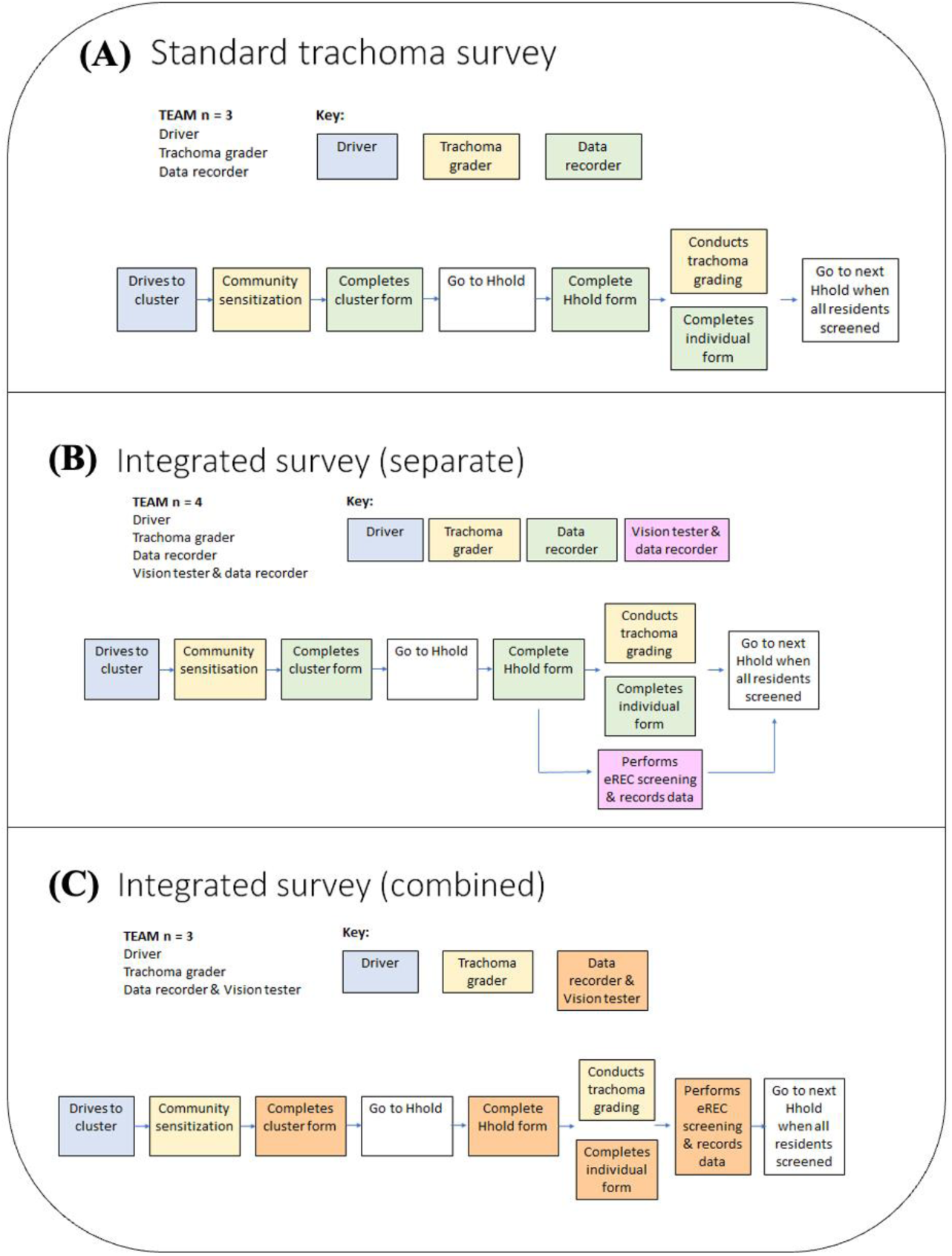
Survey options. A: Standard trachoma survey approach. Grader conducts clinical examination and recorder records the data on an Android phone. B: “Separate”: Nurse for eye health indicator assessment and data recording; separate data recorder and grader for trachoma. This is what was done for this study. C: “Combined”: Eye health indicator and trachoma data collected on one Android. Nurse records eye health indicators and then is data recorder for trachoma.

**Supplementary Figure 2.**
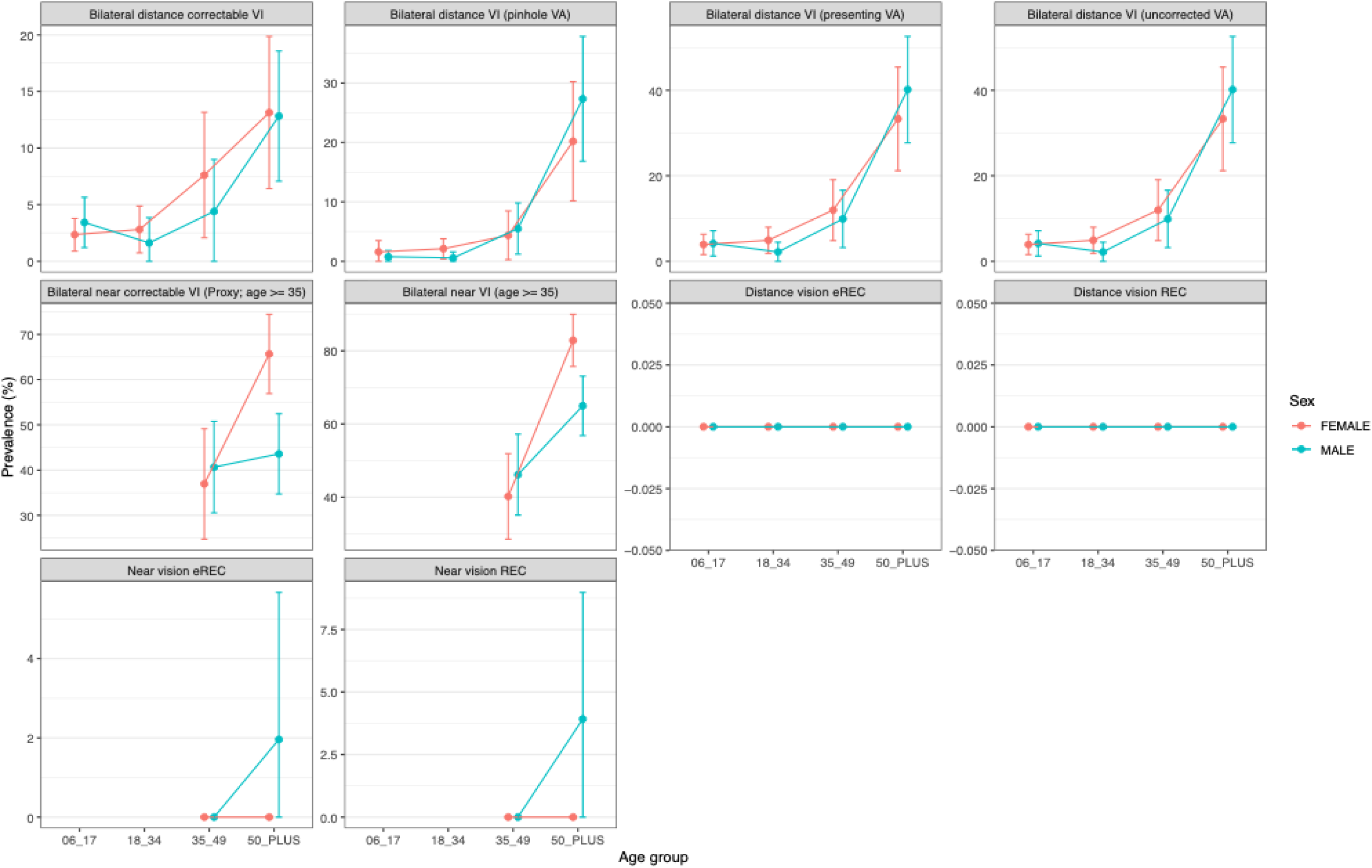
Eye Health Indicators by Sex and Age Groups – Nigeria: Standard Protocol. Age- and sex-stratified estimates of visual impairment (VI) and refractive error indicators, with prevalence (%) plotted across age groups. Points represent estimates for males and females, with error bars indicating uncertainty. VA: visual acuity; REC: refractive error coverage; eREC: effective refractive error coverage

**Supplementary Figure 3.**
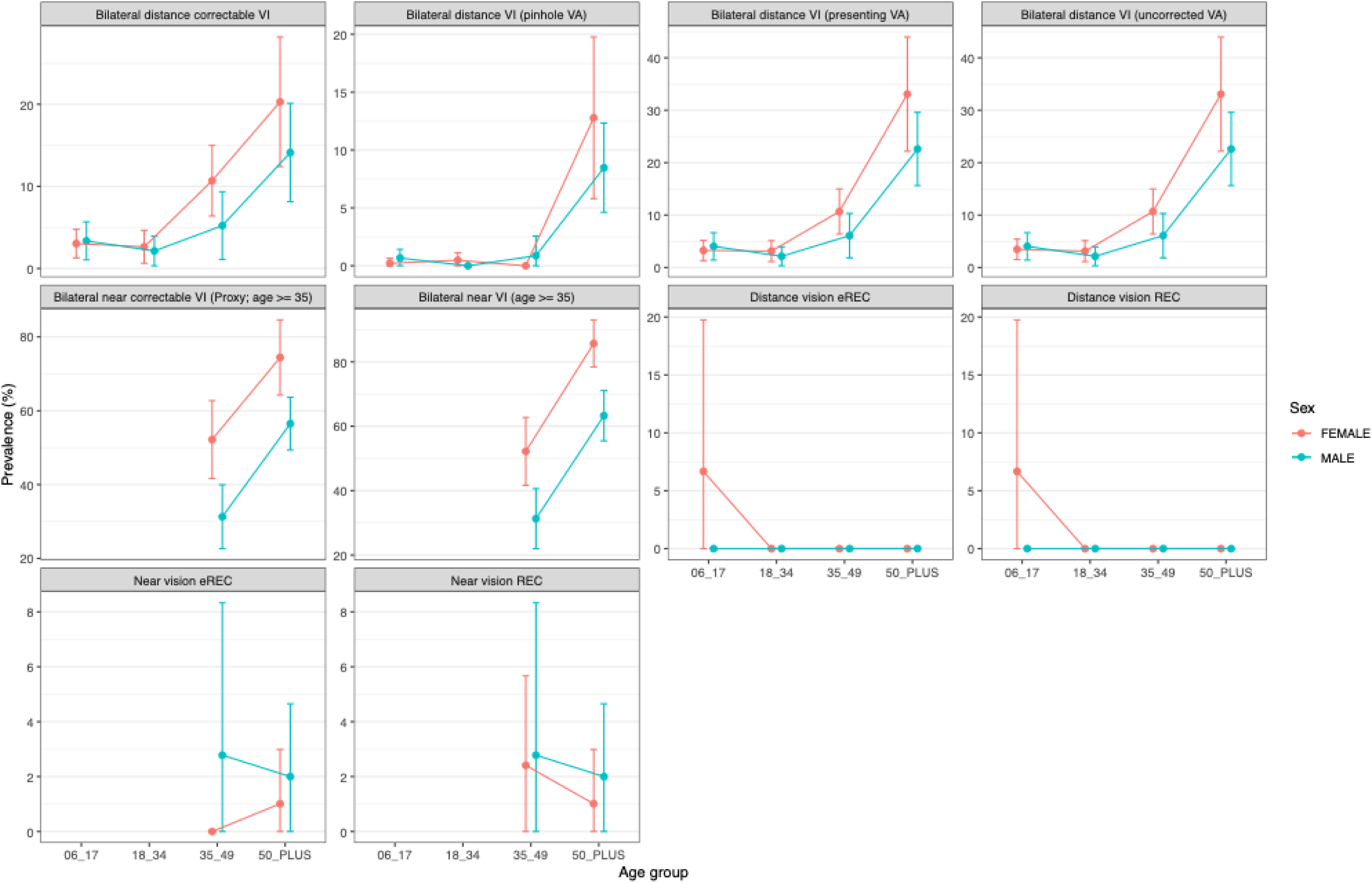
Eye Health Indicators by Sex and Age Groups – Nigeria: Minimal Protocol. Age- and sex-stratified estimates of visual impairment (VI) and refractive error indicators, with prevalence (%) plotted across age groups. Points represent estimates for males and females, with error bars indicating uncertainty. VA: visual acuity; REC: refractive error coverage; eREC: effective refractive error coverage

**Supplementary Figure 4.**
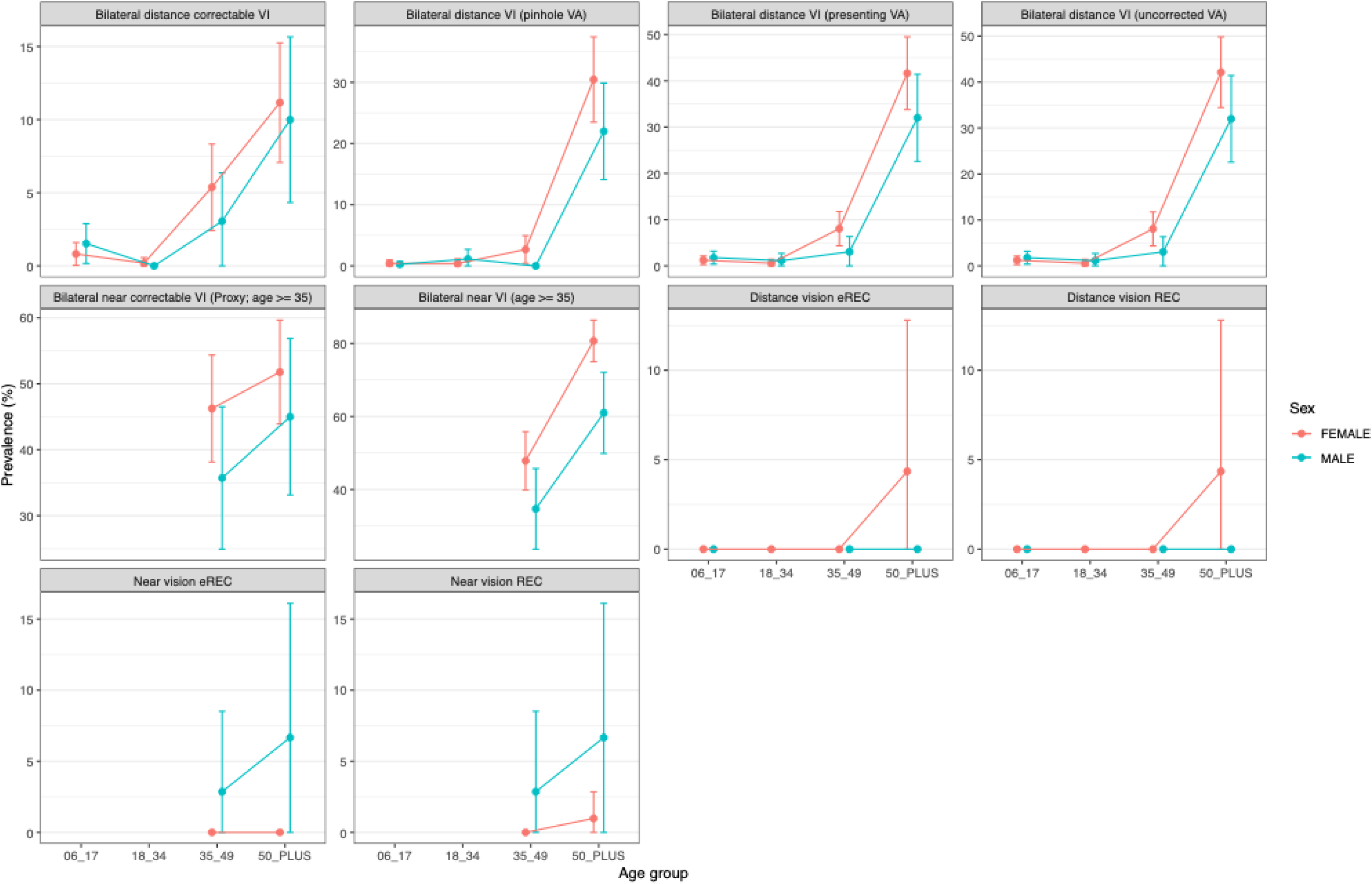
Eye Health Indicators by Sex and Age Groups – Ethiopia: Standard Protocol. Age- and sex-stratified estimates of visual impairment (VI) and refractive error indicators, with prevalence (%) plotted across age groups. Points represent estimates for males and females, with error bars indicating uncertainty. VA: visual acuity; REC: refractive error coverage; eREC: effective refractive error coverage

**Supplementary Figure 5.**
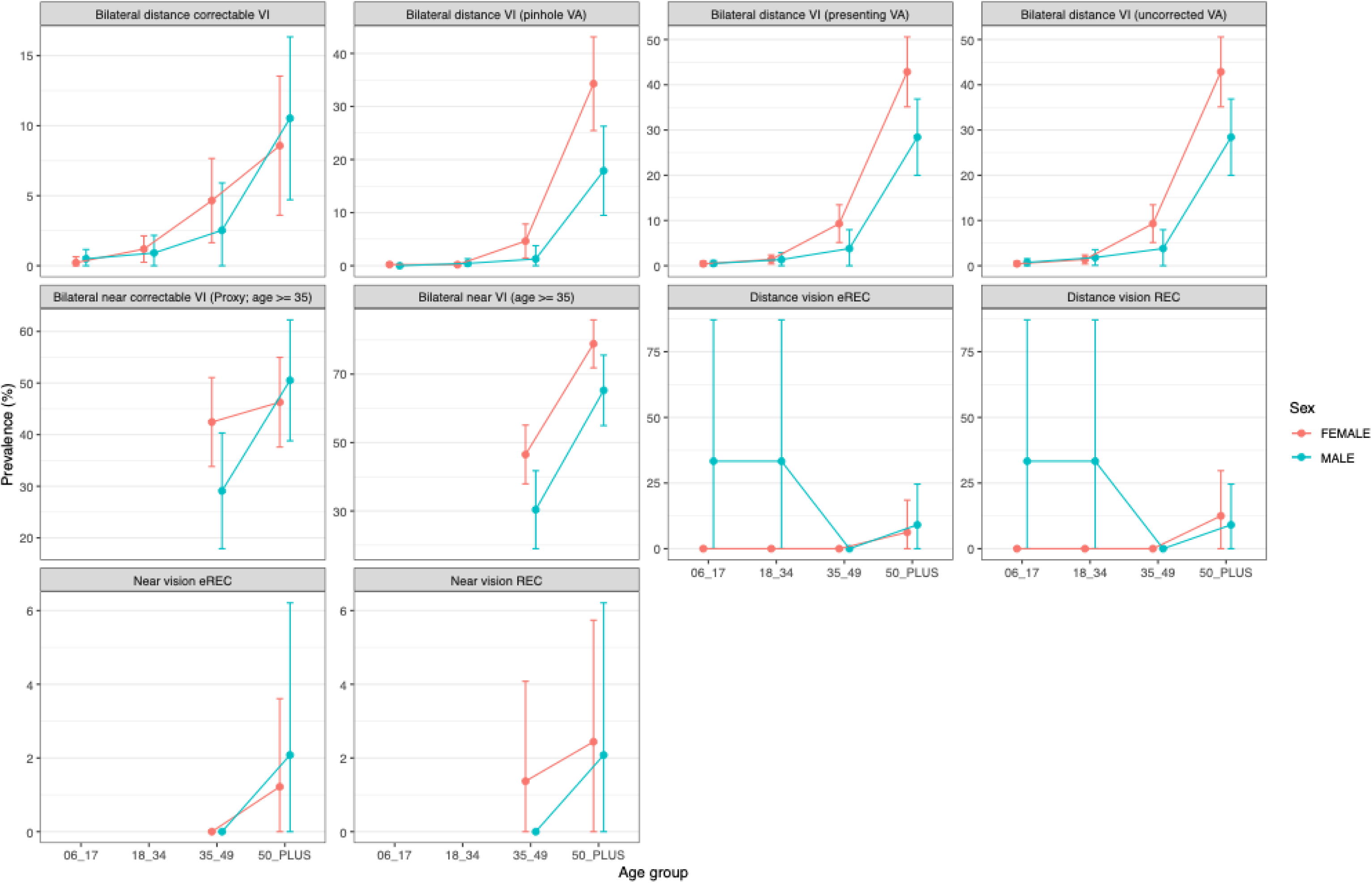
Eye Health Indicators by Sex and Age Groups – Ethiopia: Minimal Protocol. Age- and sex-stratified estimates of visual impairment (VI) and refractive error indicators, with prevalence (%) plotted across age groups. Points represent estimates for males and females, with error bars indicating uncertainty. VA: visual acuity; REC: refractive error coverage; eREC: effective refractive error coverage

**Supplementary Figure 6.**
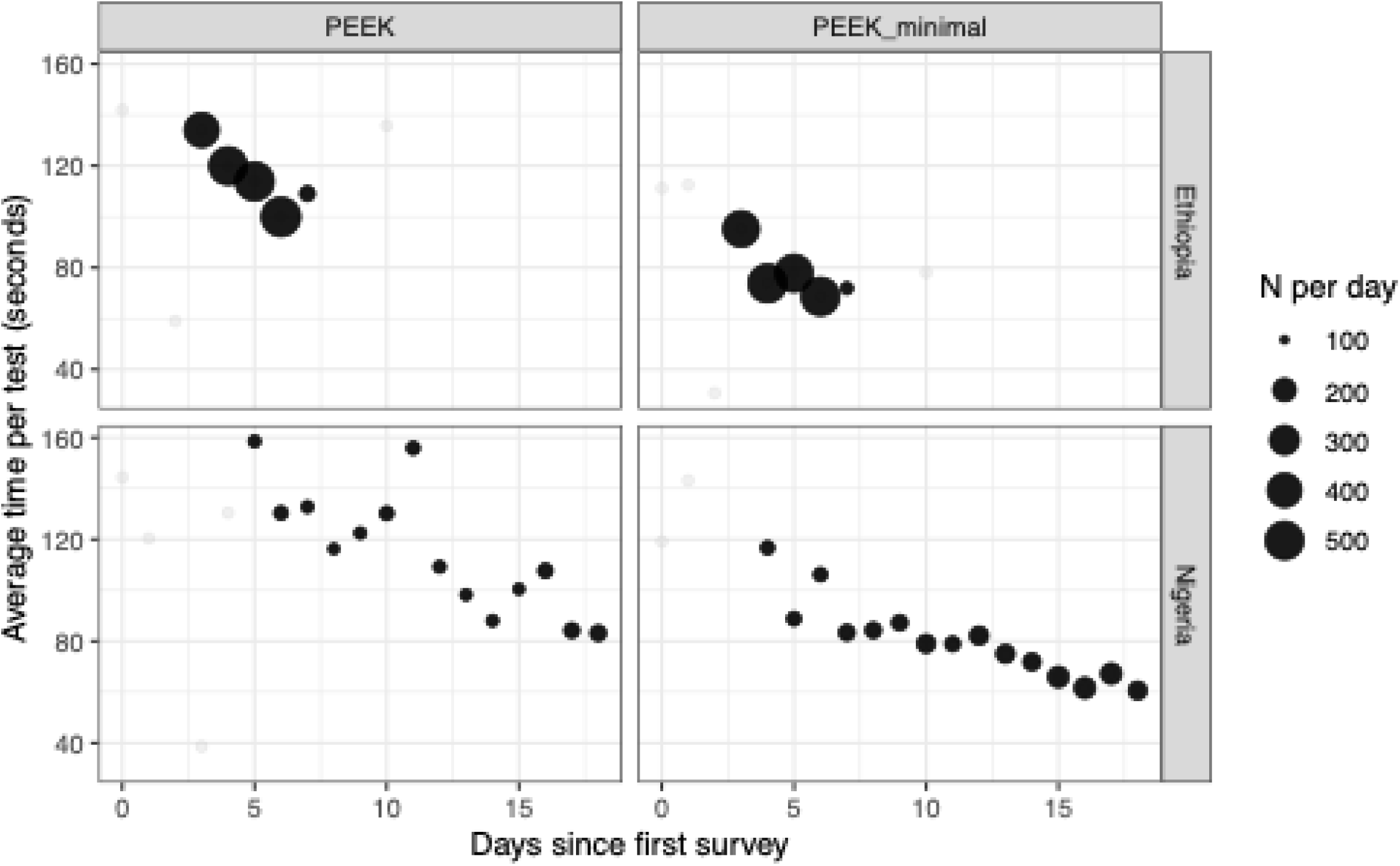
Reduction in time required for vision tests with ongoing field work observed during the study. Average time taken to complete a visual acuity test across successive days of data collection, stratified by country and protocol. Each point represents the mean time per test on a given day since the start of data collection, with point size proportional to the number of observations collected that day. Larger points therefore indicate more reliable estimates. Faint grey points show all days, including low-volume periods such as initial rollout and final mop-up, while darker points highlight days with higher sample sizes that are more representative of routine implementation.

## APPENDIX 3

### Introduction

My name is ____. I am really grateful to you for taking the time to speak with me and answer some questions. We will start off with some questions about your experiences around trachoma and eREC surveys.

This interview is to understand how well integrating data collection on vision into routine trachoma surveys works, and what could be changed to improve this method of obtaining better data on vision.

The interview will take about 20 minutes. I appreciate you spending this time with me, and sharing your thoughts and experiences.

I am going to audio record the interview to make sure that I capture all the valuable information that you share with me. I may also write things down while we’re talking so that I don’t forget anything. Participation is voluntary - you do not have to answer any question that you don’t want to, and you can choose to stop the interview at any time.

Everything you say is confidential, so please feel free to talk about your experiences and ideas. We will not record your name anywhere, and no one else will hear the tape or see the notes besides the people who are working on this research project. We may use some of what you say in reports or publications but will never use your name. If you have any questions about this study, you can ask me now, or at any time during our conversation.

The thoughts, experiences and feelings that you share are helping us to understand more about the integration of data collection on vision into routine trachoma prevalence surveys. **Do you have any questions for now?**

1. What is your role? / What was your role in this study?
2. What previous experience of trachoma and/or vision surveys do you have?

a. Follow-up: What was your role? What do you think of this experience? Would you participate again? What did you like the most? What were the difficulties encountered and how were they resolved? Was there anything in particular that you think could be modified? Provide details.
3. From your perspective, given your role, what do you think would be the advantages of integrating data collection on vision into trachoma surveys?
4. From your perspective, given your role, what do you think would be the disadvantages of integrating data collection on vision into trachoma surveys?
5. From your perspective, given your role, what do you think would be the opportunities of integrating data collection on vision into trachoma surveys?
6. From your perspective, given your role, what do you think would be the threats of integrating data collection on vision into trachoma surveys?
7. What do you consider to be the main barriers and facilitators of access to eye health services for the populations in which this study was conducted?
8. Do you know of other examples that use an existing routine survey methodology to collect data on a different disease?

a. Can you describe these examples?
b. What worked well and less well?
c. Were these one-off projects or has the integrated method been used more routinely?
9. Do you have any other feedback you want to share on integrating vision data collection into the trachoma surveys, including suggestions for the future?

For individuals involved in the fieldwork:

10. What do you think worked well during the project?
11. What do you think didn’t work well during the project?
12. What were the advantages of the different methods (standard or simplified) to measure visual acuity?
13. What were the disadvantages of the different methods (standard or simplified) to measure visual acuity?

Thank you very much for sharing your time, thoughts and experiences.

Provide space for any questions or further discussion of relevant topics.

## APPENDIX 4

World Health Organization Definitions and Calculation Methods of Distance and Near Effective Refractive Error Coverage (eREC).

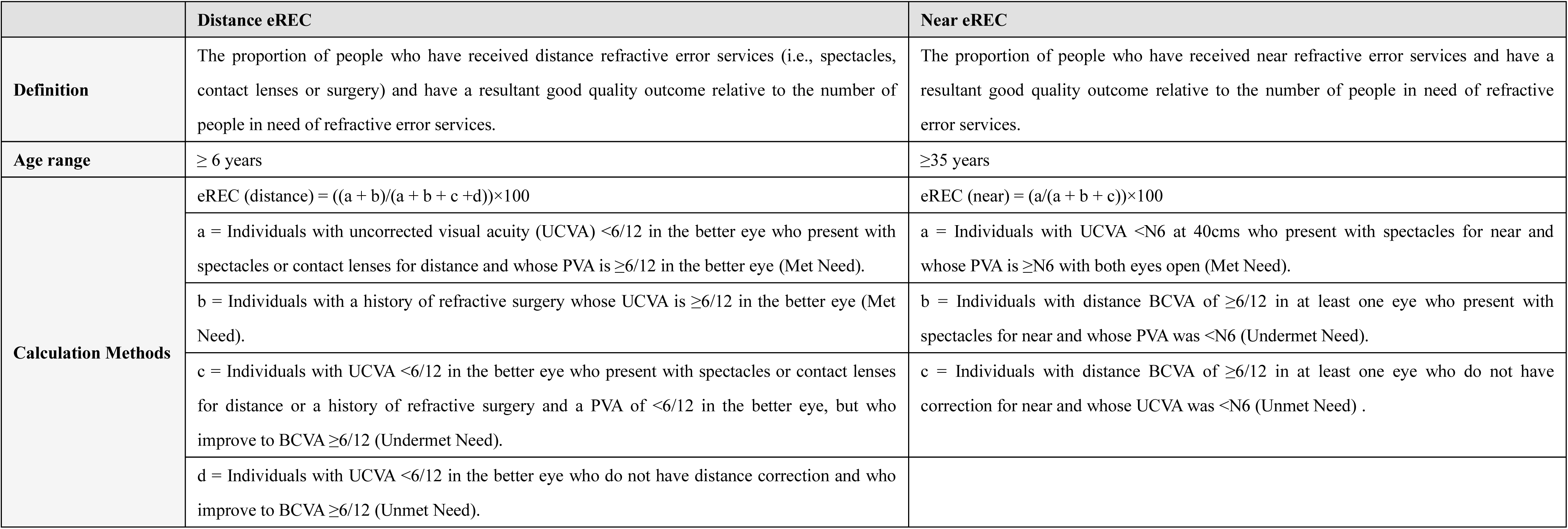

## APPENDIX 5

Comparison of time per cluster required for standalone and integrated trachoma and eye health indicator surveys.

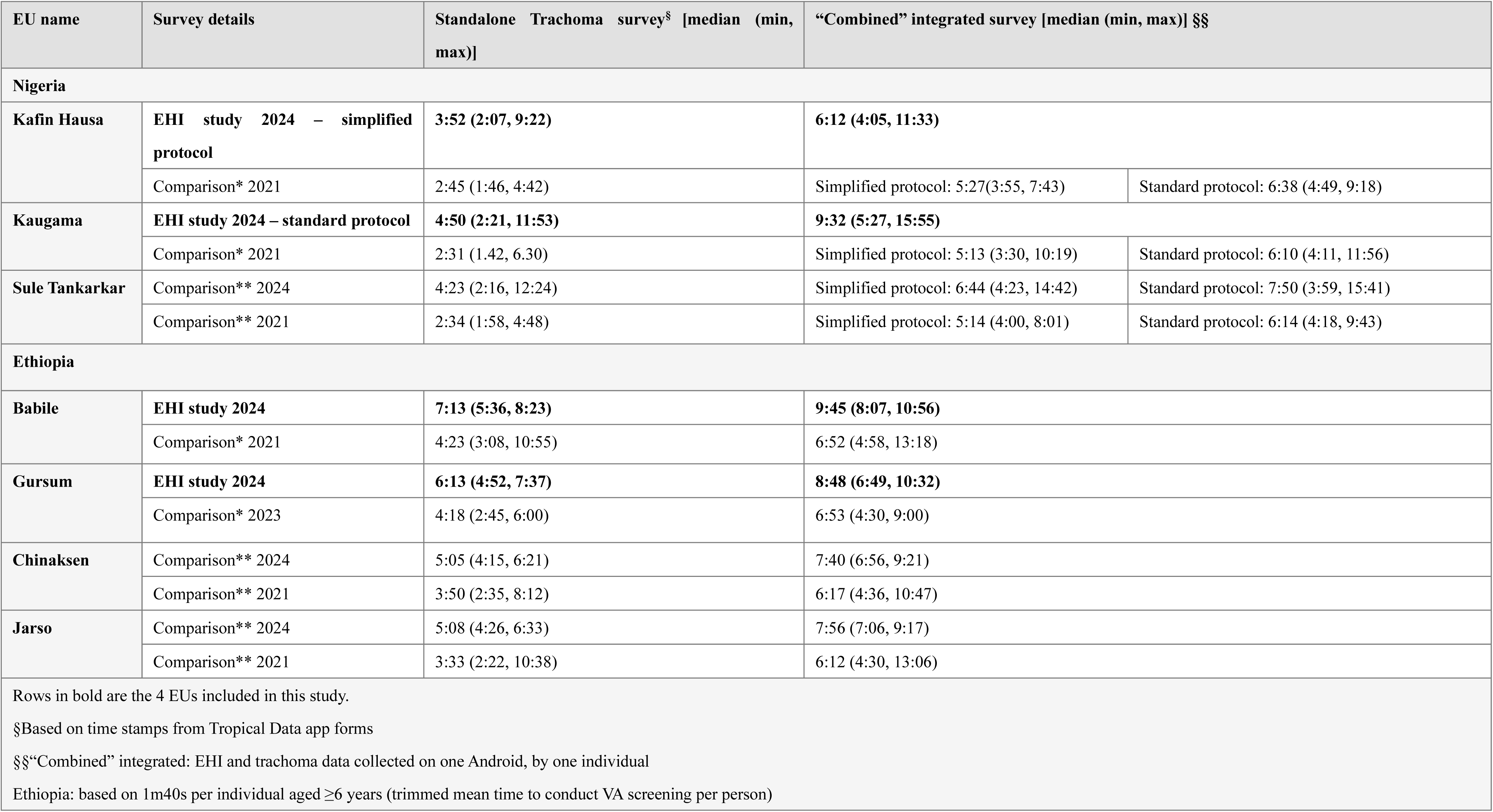
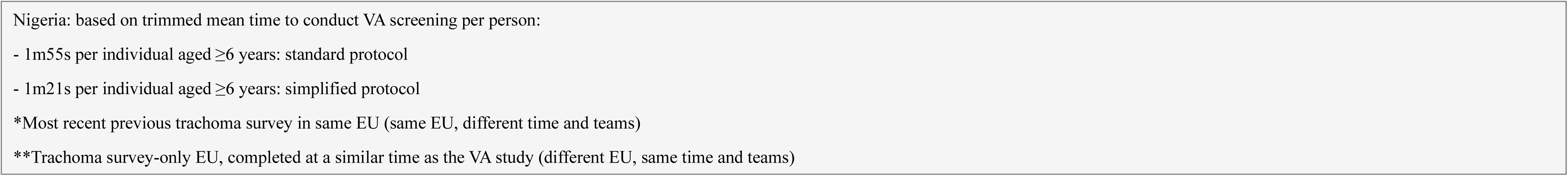

APPENDIX 5 Comparison of time per cluster required for standalone and integrated trachoma and eye health indicator surveys.
| EU name | Survey details | Standalone Trachoma survey§ [median (min, max)] | “Combined” integrated survey [median (min, max)] §§ |  |
| --- | --- | --- | --- | --- |
| Nigeria |  |  |  |  |
| Kafin Hausa | EHI study 2024 – simplified protocol | 3:52 (2:07, 9:22) | 6:12 (4:05, 11:33) |  |
|  | Comparison* 2021 | 2:45 (1:46, 4:42) | Simplified protocol: 5:27(3:55, 7:43) | Standard protocol: 6:38 (4:49, 9:18) |
| Kaugama | EHI study 2024 – standard protocol | 4:50 (2:21, 11:53) | 9:32 (5:27, 15:55) |  |
|  | Comparison* 2021 | 2:31 (1.42, 6.30) | Simplified protocol: 5:13 (3:30, 10:19) | Standard protocol: 6:10 (4:11, 11:56) |
| Sule Tankarkar | Comparison** 2024 | 4:23 (2:16, 12:24) | Simplified protocol: 6:44 (4:23, 14:42) | Standard protocol: 7:50 (3:59, 15:41) |
|  | Comparison** 2021 | 2:34 (1:58, 4:48) | Simplified protocol: 5:14 (4:00, 8:01) | Standard protocol: 6:14 (4:18, 9:43) |
| Ethiopia |  |  |  |  |
| Babile | EHI study 2024 | 7:13 (5:36, 8:23) | 9:45 (8:07, 10:56) |  |
|  | Comparison* 2021 | 4:23 (3:08, 10:55) | 6:52 (4:58, 13:18) |  |
| Gursum | EHI study 2024 | 6:13 (4:52, 7:37) | 8:48 (6:49, 10:32) |  |
|  | Comparison* 2023 | 4:18 (2:45, 6:00) | 6:53 (4:30, 9:00) |  |
| Chinaksen | Comparison** 2024 | 5:05 (4:15, 6:21) | 7:40 (6:56, 9:21) |  |
|  | Comparison** 2021 | 3:50 (2:35, 8:12) | 6:17 (4:36, 10:47) |  |
| Jarso | Comparison** 2024 | 5:08 (4:26, 6:33) | 7:56 (7:06, 9:17) |  |
|  | Comparison** 2021 | 3:33 (2:22, 10:38) | 6:12 (4:30, 13:06) |  |
| Rows in bold are the 4 EUs included in this study. |  |  |  |  |
| §Based on time stamps from Tropical Data app forms |  |  |  |  |
| §§“Combined” integrated: EHI and trachoma data collected on one Android, by one individual |  |  |  |  |
| Ethiopia: based on 1m40s per individual aged ≥6 years (trimmed mean time to conduct VA screening per person) |  |  |  |  |

Nigeria: based on trimmed mean time to conduct VA screening per person:
- 1m55s per individual aged $\geq 6$ years: standard protocol
- 1m21s per individual aged $\geq 6$ years: simplified protocol
\*Most recent previous trachoma survey in same EU (same EU, different time and teams)
\*\*Trachoma survey-only EU, completed at a similar time as the VA study (different EU, same time and teams)

## References

1. World Health Organization. World report on vision. Geneva: World Health Organization, 2019.

2. GBD 2019 Blindness and Vision Impairment Collaborators; Vision Loss Expert Group of the Global Burden of Disease Study Trends in prevalence of blindness and distance and near vision impairment over 30 years: an analysis for the Global Burden of Disease Study. Lancet Glob Health 2021; 9(2): e130–e43.

3. Hanson K, Brikci N, Erlangga D, et al. The Lancet Global Health Commission on financing primary health care: putting people at the centre. Lancet Glob Health 2022; 10(5): e715–e72.

4. World Health Organization. Eye care indicator menu (ECIM): a tool for monitoring strategies and actions for eye care provision: World Health Organization, 2022.

5. Keel S, Müller A, Block S, et al. Keeping an eye on eye care: monitoring progress towards effective coverage. Lancet Glob Health 2021; 9(10): e1460–e4.

6. Bourne RRA, Cicinelli MV, Selby DA, et al. Effective refractive error coverage in adults: a systematic review and meta-analysis of updated estimates from population-based surveys in 76 countries modelling the path towards the 2030 global target. Lancet Glob Health 2025; 13(8): e1396–e405.

7. World Health Organization. WHO Results Framework: Delivering a measurable impact in countries. 22 May 2024. https://www.who.int/publications/m/item/who-results-framework-delivering-a-measurable-impact-in-countries (accessed May 24 2026).

8. World Health Organization. Integrated people-centred eye care, including preventable vision impairment and blindness: report by the Director-General. Geneva: World Health Organization, 2021.

9. Bourne RRA, Cicinelli MV, Sedighi T, et al. Effective refractive error coverage in adults aged 50 years and older: estimates from population-based surveys in 61 countries. Lancet Glob Health 2022; 10(12): e1754–e63.

10. World Health Organization. Report of the 2030 targets on effective coverage of eye care: World Health Organization, 2022.

11. Lu F, Jiang H, Wong W, et al. Effective refractive error coverage and vision impairment among schoolchildren in Xinjiang, China. Br J Ophthalmol 2026.

12. Harding-Esch EM, Burgert-Brucker CR, Jimenez C, et al. Tropical Data: Approach and Methodology as Applied to Trachoma Prevalence Surveys. Ophthalmic Epidemiol 2023; 30(6): 544–60.

13. Ghaferi AA, Schwartz TA, Pawlik TM. STROBE Reporting Guidelines for Observational Studies. JAMA Surg 2021; 156(6): 577–8.

14. World Health Organization. Design parameters for population-based trachoma prevalence surveys. 2018. https://www.who.int/publications/i/item/who-htm-ntd-pct-2018.07 (accessed May,24 2026).

15. International Coalition for Trachoma Control. Training system for trachoma prevalence surveys. Version 4. London: International Coalition for Trachoma Control, 2019.

16. Katibeh M, Sanyam SD, Watts E, et al. Development and Validation of a Digital (Peek) Near Visual Acuity Test for Clinical Practice, Community-Based Survey, and Research. Transl Vis Sci Technol 2022; 11(12): 18.

17. Harding-Esch EM, Bakhtiari A, Boyd S, et al. Tropical Data: supporting health ministries worldwide to conduct high-quality trachoma surveys. Int Health 2025; 17(1): 1–3.

18. LSHTM Global Health Analytics. OBK. https://opendatakit.lshtm.ac.uk/ (accessed May 24 2026).

19. Zuo H, Cheng H, Lin M, et al. The effect of aging on the ciliary muscle and its potential relationship with presbyopia: a literature review. PeerJ 2024; 12: e18437.

20. Team RC. R: A language and environment for statistical computing. Vienna, 2025.

21. McCormick I. The rapid assessment of avoidable blindness (RAAB) survey methodology. Community Eye Health 2022; 35(117): 4.

22. Wilcox RR, Keselman HJ. Using Trimmed Means to Compare K Measures Corresponding to Two Independent Groups. Multivariate Behav Res 2001; 36(3): 421–44.

23. Chapman AL, Hadfield M, Chapman CJ. Qualitative research in healthcare: an introduction to grounded theory using thematic analysis. J R Coll Physicians Edinb 2015; 45(3): 201–5.

24. Fricke TR, Tahhan N, Resnikoff S, et al. Global Prevalence of Presbyopia and Vision Impairment from Uncorrected Presbyopia: Systematic Review, Meta-analysis, and Modelling. Ophthalmology 2018; 125(10): 1492–9.

25. World Health Organization. Ending the neglect to attain the sustainable development goals: a road map for neglected tropical diseases 2021–2030. Geneva: World Health Organization, 2020.

26. Renneker KK, Emerson PM, Hooper PJ, Ngondi JM. Forecasting the elimination of active trachoma: An empirical model. PLoS Negl Trop Dis 2022; 16(7): e0010563.

27. Harding-Esch EM, Brady MA, Angeles CAC, et al. Lessons from the Field: Integrated survey methodologies for neglected tropical diseases. Trans R Soc Trop Med Hyg 2021; 115(2): 124–6.

28. Fred Hollows PC. Fred Hollows: An Autobiography. 4th ed. ed. Redfern, NSW: Kerr Publishing; 1997.

29. Trotignon G, Jones I, Yasmin S, et al. Compliance with spectacle use among schoolchildren in a school health programme in Islamabad, Pakistan. Int Health 2025; 17(Supplement_1): i21–i9.

30. Wu Y, Keel S, Carneiro VLA, et al. Real-world application of a smartphone-based visual acuity test (WHOeyes) with automatic distance calibration. Br J Ophthalmol 2024; 108(11): 1613–20.

